# Immunogenicity of SARS-CoV-2 infection and Ad26.CoV2.S vaccination in people living with HIV

**DOI:** 10.1101/2021.10.08.21264519

**Authors:** Khadija Khan, Gila Lustig, Mallory Bernstein, Derseree Archary, Sandile Cele, Farina Karim, Muneerah Smith, Yashica Ganga, Zesuliwe Jule, Kajal Reedoy, Yoliswa Miya, Ntombifuthi Mthabela, COMMIT-KZN Team, Richard Lessells, Tulio de Oliveira, Bernadett I. Gosnell, Salim Abdool Karim, Nigel Garrett, Willem Hanekom, Linda Gail Bekker, Glenda Gray, Jonathan M. Blackburn, Mahomed-Yunus S. Moosa, Alex Sigal

## Abstract

**Background:** People living with HIV (PLWH) have been reported to have an increased risk of more severe COVID-19 disease outcome and an increased risk of death relative to HIV-uninfected individuals. Here we assessed the ability of the Johnson and Johnson Ad26.CoV2.S vaccine to elicit neutralizing antibodies to the Delta variant in PLWH relative to HIV-uninfected individuals. We also compared the neutralization after vaccination to neutralization elicited by SARS-CoV-2 infection only in HIV-uninfected, suppressed HIV PLWH, and PLWH with detectable HIV viremia.

**Methods:** We enrolled 26 PLWH and 73 HIV-uninfected participants from the SISONKE phase 3b open label South African clinical trial of the Ad26.CoV2.S vaccine in health care workers (HCW). Enrollment was a median 56 days (range 19-98 days) post-vaccination and PLWH in this group had well controlled HIV infection. We also enrolled unvaccinated participants previously infected with SARS-CoV-2. This group consisted of 34 PLWH and 28 HIV-uninfected individuals. 10 of the 34 (29%) SARS-CoV-2 infected only PLWH had detectable HIV viremia. We used records of a positive SARS-CoV-2 qPCR result, or when a positive result was absent, testing for SARS-CoV-2 nucleocapsid antibodies, to determine which vaccinated participants were SARS-CoV-2 infected prior to vaccination. Neutralization capacity was assessed using participant plasma in a live virus neutralization assay of the Delta SARS-CoV-2 variant currently dominating infections in South Africa. This study was approved by the Biomedical Research Ethics Committee at the University of KwaZulu–Natal (reference BREC/00001275/2020).

**Findings:** The majority (68%) of Ad26.CoV2.S vaccinated HCW were found to be previously infected with SARS-CoV-2. In this group, Delta variant neutralization was 9-fold higher compared to the infected only group (GMT=306 versus 36, p<0.0001) and 26-fold higher relative to the vaccinated only group (GMT=12, p<0.0001). No significant difference in Delta variant neutralization capacity was observed in vaccinated and previously SARS-CoV-2 infected PLWH relative to vaccinated and previously SARS-CoV-2 infected, HIV-uninfected participants (GMT=307 for HIV-uninfected, 300 for PLWH, p=0.95). SARS-CoV-2 infected, unvaccinated PLWH showed 7-fold reduced neutralization of the Delta variant relative to HIV-uninfected participants (GMT=105 for HIV-uninfected, 15 for PLWH, p=0.001). There was a higher frequency of non-responders in PLWH relative to HIV-uninfected participants in the SARS-CoV-2 infected unvaccinated group (27% versus 0%, p=0.0029) and 60% of HIV viremic versus 13% of HIV suppressed PLWH were non-responders (p=0.0088). In contrast, the frequency of non-responders was low in the vaccinated/infected group, and similar between HIV-uninfected and PLWH. Vaccinated only participants showed a low neutralization of the Delta variant, with a stronger response in PLWH (GMT=6 for HIV-uninfected, 73 for PLWH, p=0.02).

**Interpretation:** The neutralization response of the Delta variant following Ad26.CoV2.S vaccination in PLWH with well controlled HIV was not inferior to HIV-uninfected study participants. In SARS-CoV-2 infected and non-vaccinated participants, the presence of HIV infection reduced the neutralization response to SARS-CoV-2 infection, and this effect was strongest in PLWH with detectable HIV viremia

**Funding:** South African Medical Research Council, The Bill & Melinda Gates Foundation.

## Introduction

South Africa has a high burden of HIV infection (1) and recent studies observed Covid-19 disease severity (2) and mortality risk (3) are increased among people living with HIV (PLWH). HIV interferes with protective vaccination against multiple pathogens, usually through the decreased effectiveness of the antibody response (4-8). HIV infection reduces the number of CD4 T cells (9), the primary target cells of the virus in different compartments (10), and reduced CD4 T cell numbers correlate with reduced concentrations of antibodies to SARS-CoV-2 (11).

The effects of HIV status on vaccine efficacy are still being determined. While numbers the numbers of PLWH participants were very small, there was no efficacy of the Novavax NVX-CoV2373 vaccine in PLWH (12). SARS-CoV-2 vaccine efficacy may also be reduced by infection with a SARS-CoV-2 variant. Infection with the Beta variant (13-15) was associated with a drop in neutralization capacity of the AstraZeneca ChAdOx vaccine and this led to a loss of vaccine efficacy in South Africa (16). The effect of HIV status on the protection mediated by the Johnson and Johnson Ad26.CoV2.S vaccine is yet unknown.

SARS-CoV-2 neutralization by antibodies correlates with SARS-CoV-2 vaccine efficacy (17) and may be a rapid preliminary predictor of vaccine efficacy where efficacy data is not yet available. Two studies assessing immunogenicity of the ChAdOx1 nCoV-19 of the chimpanzee adenovirus vectored vaccine observed comparable anti-SARS-CoV-2 spike receptor binding domain (RBD) antibody levels and similar enhancement of SARS-CoV-2 neutralization after vaccination of PLWH previously exposed to SARS-CoV-2 relative to HIV-uninfected participants (18, 19). Pfizer-BNT162b2 mRNA vaccination of PLWH with well controlled HIV and CD4 T cell counts of >200 resulted in similar SARS-CoV-2 antibody levels and neutralization relative to HIV-uninfected individuals (20-24). This was also the case in a recent study of the Sinopharm inactivated SARS-CoV-2 vaccine (25). While vaccine elicited neutralization in PLWH vaccinated with Ad26.CoV2.S has previously not been reported, data from HIV-uninfected participants in the SISONKE phase 3b (26) open label South African clinical trial of the Ad26.CoV2.S vaccine in health care workers (HCW) showed moderate neutralization in vaccinated participants which was enhanced by previous infection (27).

Here we investigated whether the Johnson and Johnson Ad26.CoV2.S vaccine elicits a comparable neutralizing response to the Delta variant (13) in PLWH relative to HIV-uninfected study participants using a live virus neutralization assay. We compared the results to SARS-CoV-2 infected, unvaccinated PLWH and HIV-uninfected participants. We observed that well controlled HIV infection does not reduce the Ad26.CoV2.S vaccine elicited neutralization response. In SARS-CoV-2 infected only participants, we observed that HIV infection did interfere with the neutralization response to SARS-CoV-2 and interference was strongest in HIV viremic PLWH.

## Results

We tested SARS-CoV-2 neutralization in Ad26.CoV2.S vaccinated HIV-uninfected and PLWH participants from the SISONKE trial. We used a live virus neutralization assay of the Delta variant, dominant in the latest, third infection wave in South Africa (28). We categorized participants into vaccinated only, previously infected and vaccinated, and SARS-CoV-2 infected unvaccinated. The time post-infection of samples from the infection only group were matched as closely as possible to the median time post-infection in the infected/vaccinated group (range 6-10 months, Table 1).

**TABLE 1:** Study participant characteristics

|  | *Infected unvaccinated* |  |  | *Infected and vaccinated* |  |  | *Vaccinated only* |  |  |
| --- | --- | --- | --- | --- | --- | --- | --- | --- | --- |
|  | All | HIV- | HIV+ | All | HIV- | HIV+ | All | HIV- | HIV+ |
| **Number participants** | 62 | 28<br>(45.2%) | 34<br>(54.8%) | 67 | 49<br>(73.1%) | 18<br>(26.9%) | 32 | 24<br>(75.0%) | 8<br>(25.0%) |
| **Age years** | 44<br>(39 - 57) | 57<br>(46 - 64) | 41<br>(35 - 45) | 46<br>(40 - 52) | 46<br>(40 - 52) | 47<br>(42 - 51) | 45<br>(39 - 52) | 48<br>(42 - 55) | 39<br>(36 - 42) |
| **Days post-infection** | 188<br>(120 - 278) | 192<br>(108 - 279) | 187<br>(122 - 277) | 235<br>(141 - 306) | 230<br>(134 - 303) | 304<br>(187 - 333) | - | - | - |
| **Days post vaccination** | - | - | - | 48<br>(34 - 81) | 48<br>(34 - 80) | 51<br>(34 - 86) | 74<br>(50 - 84) | 74<br>(44 - 85) | 74<br>(61 - 82) |
| **Male sex** | 12 (19.4%) | 5 (17.9%) | 7 (20.6%) | 2 (3.0%) | 2 (4.1%) | 0 (0.0%) | 1 (3.1%) | 0 (0.0%) | 1 (12.5%) |
| **Number HIV viremic** | - | - | 10<br>(29.4%) | - | - | 1<br>(5.6%) | - | - | 0<br>(0.0%) |
| **HIV viral load** | - | - | 3060<br>(1224 - 30160) | - | - | 3219 | - | - | - |
| **Years ART** | - | - | 11<br>(5 - 15) | - | - | 7<br>(5 - 12) | - | - | 5<br>(4 - 11) |
| **CD4 count cells/μL** | 792<br>(513 - 1027) | 991<br>(807 - 1179) | 581<br>(328 - 794) | 967<br>(784 - 1325) | 1033<br>(877 - 1424) | 852<br>(730 - 1184) | 1199<br>(853 - 1368) | 1215<br>(1101 - 1413) | 735<br>(458 - 863) |
| **CD4/CD8 ratio** | 1.1<br>(0.7 - 1.2) | 1.6<br>(1.3 - 2.1) | 0.8<br>(0.4 - 1.1) | 1.6<br>(1.1 - 2.2) | 1.7<br>(1.4 - 2.3) | 1.1<br>(0.8 - 1.2) | 1.8<br>(1.2 - 2.1) | 1.9<br>(1.2 - 2.3) | 1.1<br>(0.4 - 1.2) |

All values are median (IQR). Fraction HIV viremic is number PLWH with HIV RNA >40 copies/ml of total PLWH. Median HIV viral load is for HIV viremic participants only

We used a record of a SARS-CoV-2 positive qPCR test as an indication of previous SARS-CoV-2 infection for all SARS-CoV-2 infected unvaccinated participants and the vaccinated participants where such a record was available. To account for asymptomatic or unreported SARS-CoV-2 infection in vaccinated participants, we tested for the presence or SARS-CoV-2 nucleocapsid protein antibodies, which are made against the nucleocapsid protein produced in infection but not delivered by Ad26.CoV2.S vaccination. Therefore, a participant was considered previously infected if either nucleocapsid antibodies were detected (Supplementary Figure 1) or a previous positive qPCR for SARS-CoV-2 existed. Of the vaccinated participants, 68% were found to be previously infected with SARS-CoV-2 (Supplementary Figure 1).

The results showed that the SARS-CoV-2 infected and non-vaccinated participants had low but detectable SARS-CoV-2 Delta variant neutralization measured as PRNT50, the inverse of the dilution required for 50% neutralization (Figure 1). Neutralization was significantly higher in the group receiving Ad26.CoV2.S vaccination relative to the infected unvaccinated group (geometric mean titer (GMT) of 306 versus 36, a 9-fold increase, p<0.0001). Neutralization in the vaccinated/infected group was also 26-fold higher than in the vaccinated only group (GMT=12, p<0.0001), although the PRNT50 in the latter was below the lowest dilution tested and therefore extrapolated.

**Figure 1:**
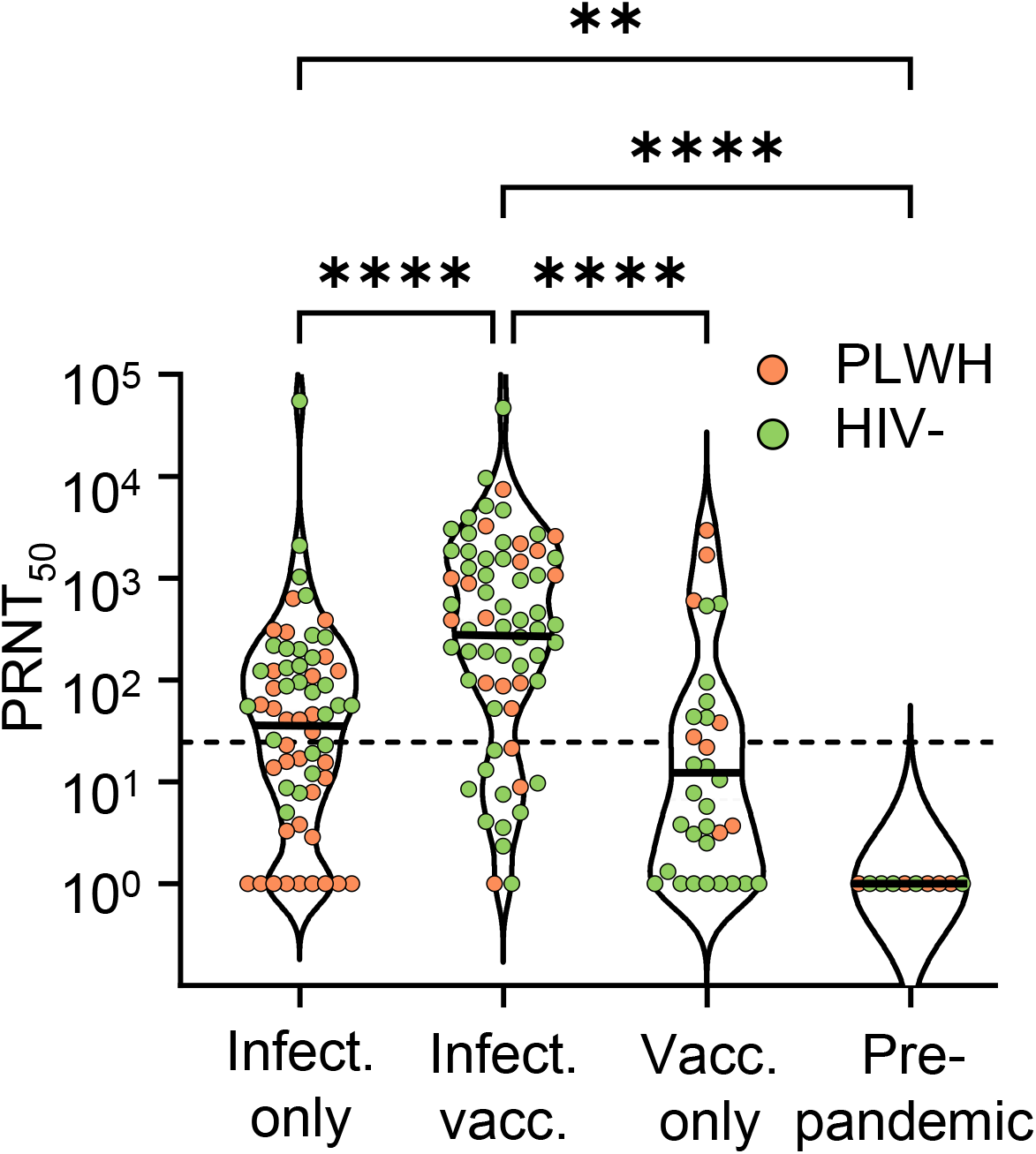
Effect of previous SARS-CoV-2 exposure on neutralization capacity elicited by Ad26.CoV2.S. Violin plots of neutralization capacity of Delta variant as PRNT50 in SARS-CoV-2 infected unvaccinated, infected and vaccinated, vaccinated only, and pre-pandemic participants. PLWH are represented by orange points and HIV negative participants by green points. Horizontal lines represent GMT. Participant numbers per category were n=62 (34 PLWH, 28 HIV-) for infected unvaccinated, n=67 (18 PLWH, 49 HIV-) for infected vaccinated, and n=32 (8 PLWH, 24 HIV-) for vaccinated only. p-values are ** <0.01, **** < 0.0001 as determined by the Kruskal-Wallis test with Dunn multiple hypothesis correction. Dashed horizontal line denotes most concentrated plasma tested.

In the infected unvaccinated group, neutralization of the Delta variant was 7-fold lower in PLWH relative to HIV negative participants (Figure 2A, GMT=105 for HIV-uninfected, 15 for PLWH, p=0.001). In contrast, there was no significant difference in vaccine elicited neutralization in PLWH versus HIV-uninfected participants who received the vaccine following SARS-CoV-2 infection (Figure 2B). In vaccinated only participants, PLWH seemed to have a stronger vaccine elicited neutralization with borderline significance (Figure 2C, GMT=6 for HIV negative, 73 PLWH, p=0.019).

**Figure 2:**
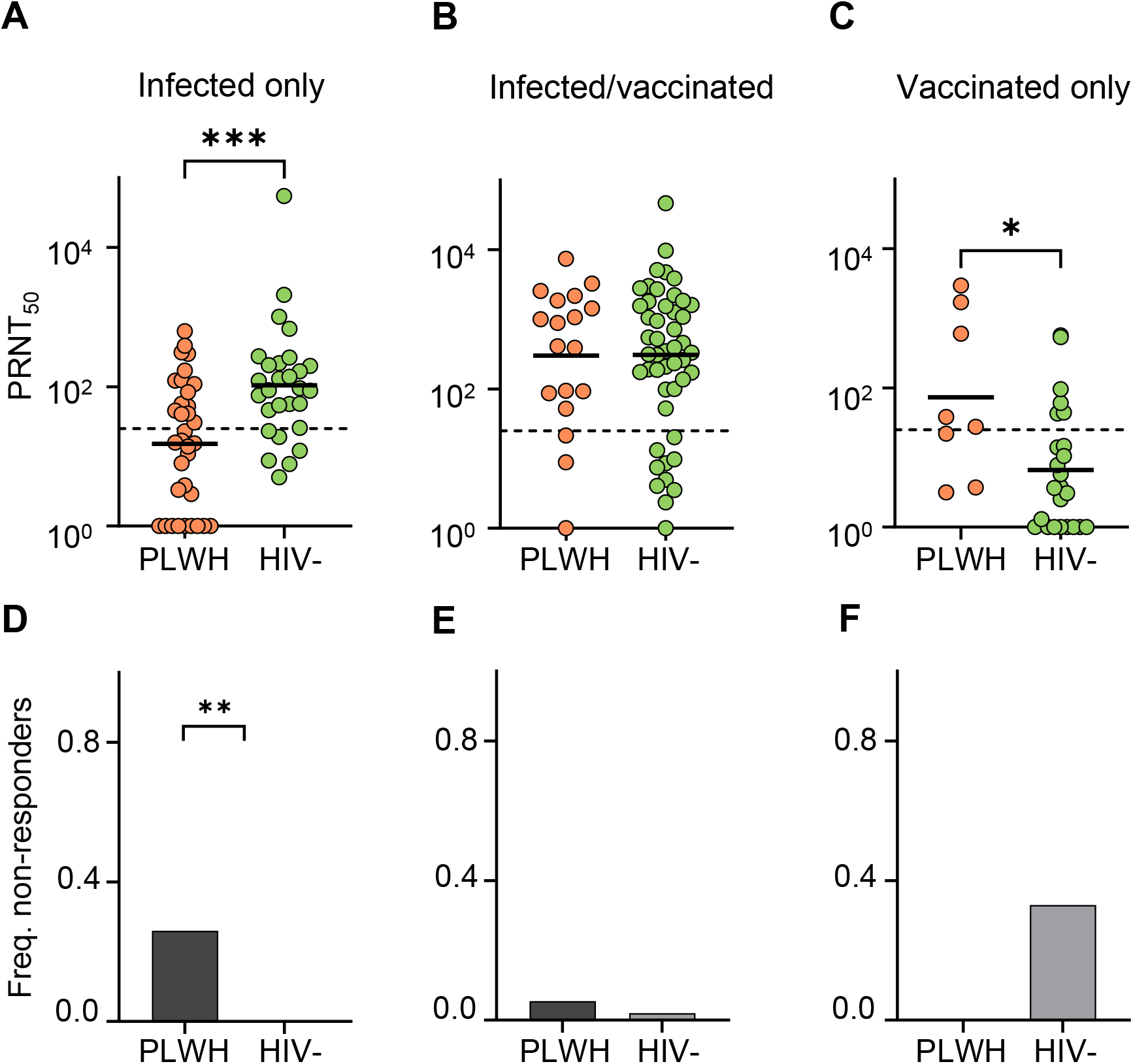
Effect of HIV status on neutralization capacity elicited by Ad26.CoV2.S. (A) Neutralization capacity as PRNT50 for Delta variant neutralization in SARS-CoV-2 infected unvaccinated (A), infected and vaccinated (B), and vaccinated only (C) participants. Solid horizontal lines represent GMT and dashed horizontal lines represent most concentrated plasma used. (D-F) frequency of participants with no detectable Delta variant neutralization (non-responders) in SARS-CoV-2 infected unvaccinated (D), infected and vaccinated (E), and vaccinated only (F) participants. p-values are * <0.05, ** <0.01, *** < 0.001, as determined for (A-C) by the Mann–Whitney U test and for (D-F) by Fisher’s exact test.

We next examined each group for non-responders, defined as no detectable neutralization of the Delta variant in the LVNA (PRNT50=1 in the graphs). The infected unvaccinated PLWH showed a frequency of 26.5% of non-responders while there were no non-responders in HIV-uninfected participants (Figure 2D). In contrast, the frequency of non-responders was only 5.6% in PLWH and 2.0% in HIV-uninfected in the vaccinated/SARS-CoV-2 infected group. The difference between PLWH and HIV-uninfected participants was not significant (p=0.47). In the vaccinated only group, there were 33.3% non-responders in the HIV-uninfected group and none in PLWH, but the difference was non-significant (p=0.082).

We next determined the effect of HIV suppression in the SARS-CoV-2 infected unvaccinated group. In this group, 29.4% of PLWH participants had detectable HIV viremia (Table 1), compared to 5.6% in the infected/vaccinated group and none in the vaccinated only group (therefore, the numbers of HIV viremic participants in the latter two groups were too small for analysis). The frequency of non-responders in the HIV viremic subset was 60.0% while it was 13.0% in the HIV suppressed subset (Figure 3A, p=0.0088). There was also a lower PRNT50 GMT (6 in HIV viremic versus 22 in suppressed) but this was non-significant (Figure 3B, p=0.13).

**Figure 3:**
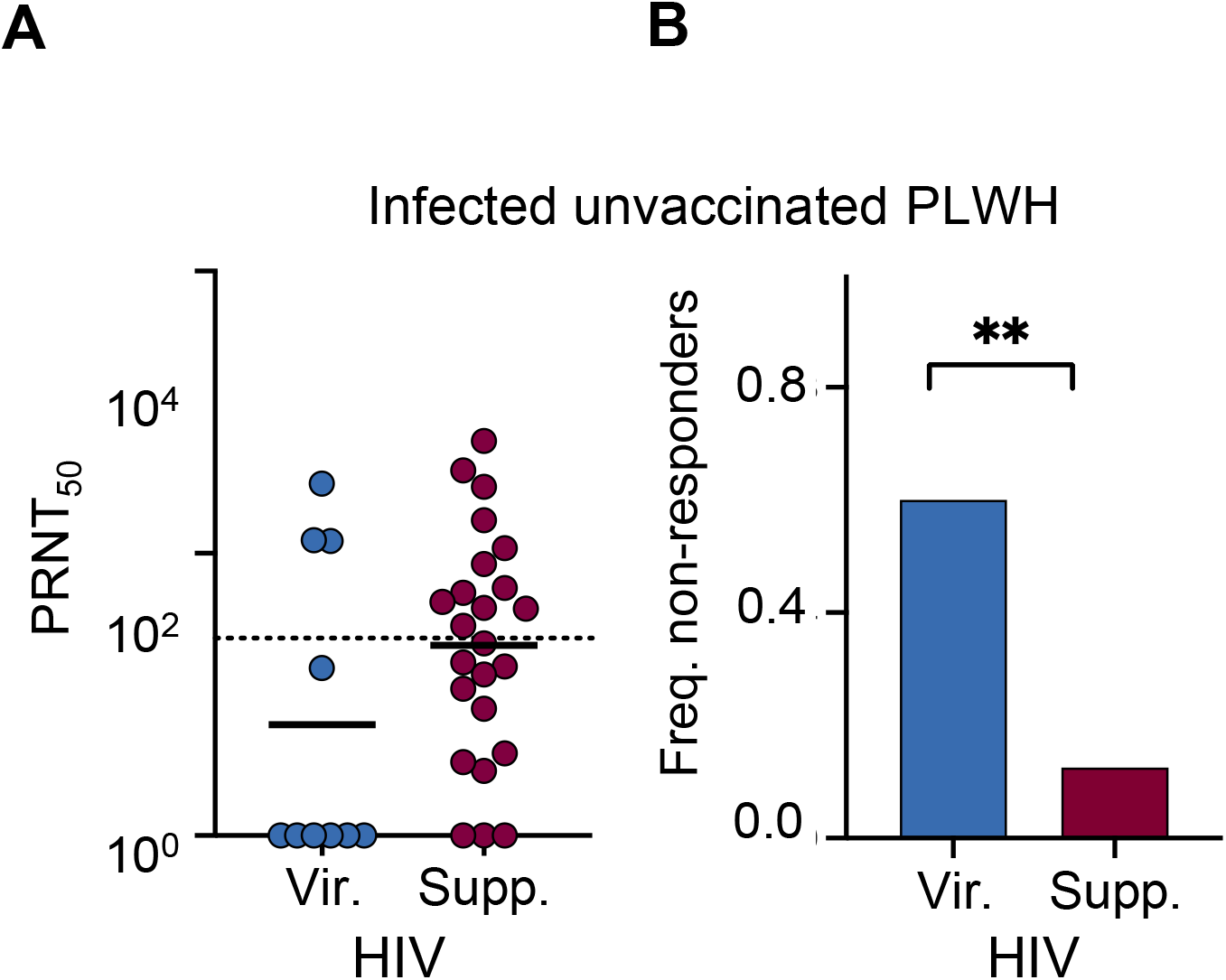
Effect of HIV viremia on neutralization capacity in infected unvaccinated participants. (A) Neutralization capacity as PRNT50 for Delta variant neutralization in SARS-CoV-2 infected unvaccinated HIV viremic (n=10) versus HIV suppressed (n=24) participants. Dashed horizontal line represents most concentrated plasma used. (B) Frequency of non-responders in (A). p-values are p=0.13 for (A) by the Mann–Whitney U test and p=0.0088 for (B) by Fisher’s exact test.

PLWH were almost all HIV suppressed in the vaccinated groups. We therefore compared the suppressed HIV PLWH in the SARS-CoV-2 infected unvaccinated group to HIV-uninfected participants. We observed that there was still lower neutralization of the Delta variant in SARS-CoV-2 infected only HIV suppressed PLWH relative to HIV-uninfected participants (Figure S2A), although the difference in the fraction of non-responders became non-significant (Figure S2B).

CD4 T cell count may be an important determinant of the immune response. The CD4 count was significantly lower in the infected unvaccinated group relative to the vaccinated only and vaccinated infected groups (Figure S3). The CD4 count was lower in PLWH relative to HIV-uninfected participants in all groups (Figure S3). In the infected only group, there was a significant correlation between higher CD4 count and higher neutralization (r=0.36, p=0.0045, Figure 4A), although this correlation was closely associated with HIV status, with lower CD4 counts measured in PLWH. There were no significant correlations between CD4 T cell count and neutralization in the infected vaccinated or vaccinated only groups (Figure 4B-C).

**Figure 4:**
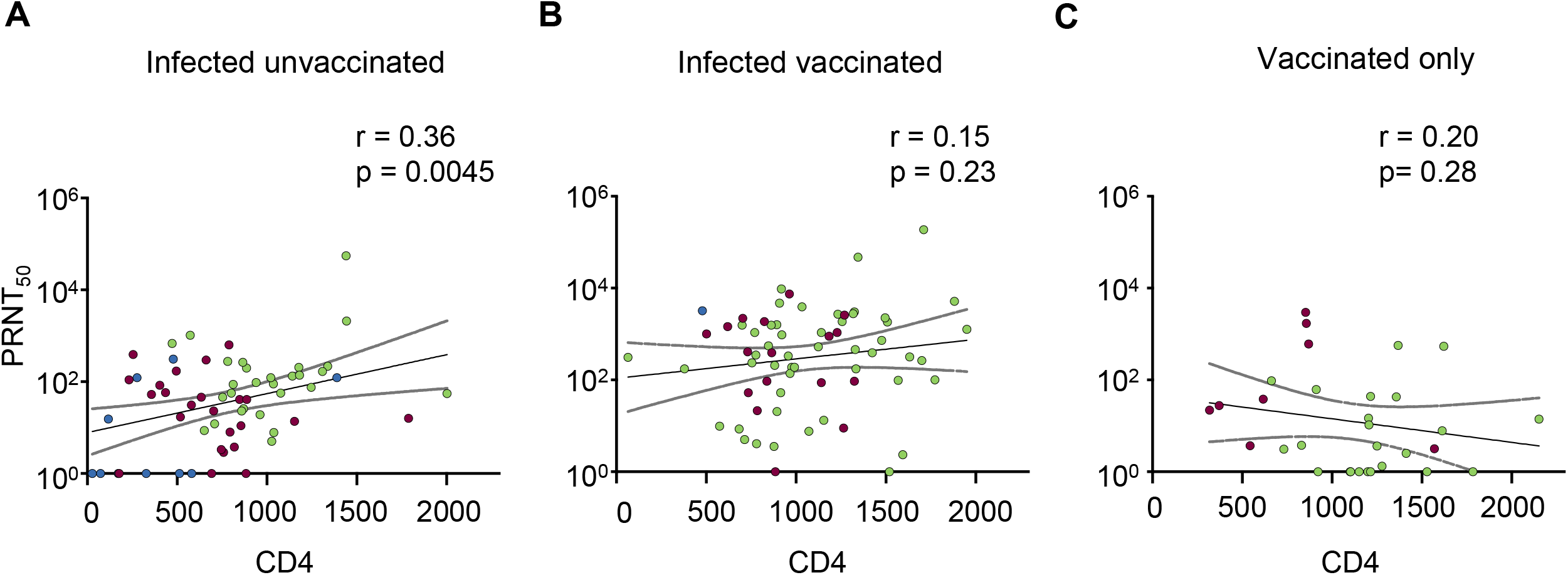
Correlation between CD4 count and neutralization capacity. Pearson correlation of PRNT50 versus CD4 count in infected only (A), infected and vaccinated (B), and vaccinated only (C). Solid lines represent linear regression and dashed lines represent 95% confidence intervals. r is the Pearson correlation coefficient. Green points are HIV-uninfected participants, purple points are PLWH with suppressed HIV viremia, and blue points are HIV viremic PLWH.

## Discussion

Our results are consistent with a non-compromised neutralization response to Ad26.CoV2.S vaccination in PLWH. We note that the vaccinated HCW PLWH tested in our study showed well controlled HIV infection and relatively high CD4 counts. The higher neutralization in vaccinated only PLWH relative to HIV-uninfected participants was surprising. However, the number of participants in the comparison was small, there was a wide dispersion in PRNT50 values, and the vaccinated only PLWH were younger, perhaps accounting for the better response (29). As was previously demonstrated, previous SARS-CoV-2 infection enhanced the neutralization response to Ad26.CoV2.S vaccination (27). We found this to be the case in PLWH as well as HIV-uninfected participants.

The effect of HIV status in both the vaccinated only and vaccinated infected groups contrasts with the infected unvaccinated group, which showed a deleterious effect of HIV infection on neutralization of the Delta variant and an increased number of non-responders, especially among PLWH with detectable HIV viremia. However, even in HIV suppressed PLWH, the neutralization response to Delta variant infection was lower.

SARS-CoV-2 infected unvaccinated participants were the only group where a moderate but significant correlation between CD4 T cell count and Delta neutralization was detected, possibly indicating that infection elicited immunity may be more sensitive than vaccination to HIV mediated changes such as a drop in the number of CD4 T cells. However, the time post-infection which was used (about 6 months) was considerably longer than time post-vaccination (about 2 months) and may be more influenced by the durability of the neutralization response. Therefore, it is yet unclear if uncontrolled HIV viremia or low CD4 count may decrease neutralization capacity after Ad26.CoV2.S vaccination, particularly at longer timepoints post-vaccination. Overall, the results indicate that vaccination with Ad26.CoV2.S has a benefit in terms of conferring SARS-CoV-2 neutralization capacity in PLWH from South Africa with well suppressed HIV infection.

## Methods

### Ethical statement

Nasopharyngeal and oropharyngeal swab samples and plasma samples were obtained from vaccinees and adults with PCR-confirmed SARS-CoV-2 infection. These participants are enrolled in a prospective cohort study approved by the Biomedical Research Ethics Committee at the University of KwaZulu–Natal (reference BREC/00001275/2020).

### Cells

Vero E6 cells (ATCC CRL-1586, obtained from Cellonex in South Africa) were propagated in complete DMEM with 10% fetal bovine serum (Hylone) containing 1% each of HEPES, sodium pyruvate, L-glutamine and nonessential amino acids (Sigma-Aldrich). Vero E6 cells were passaged every 3–4 days. The H1299-E3 cell line for first-passage SARS-CoV-2 expansion, derived as described in (15), was propagated in complete RPMI with 10% fetal bovine serum containing 1% each of HEPES, sodium pyruvate, L-glutamine and nonessential amino acids. H1299 cells were passaged every second day. Cell lines have not been authenticated. The cell lines have been tested for mycoplasma contamination and are mycoplasma negative.

### Virus expansion

All work with live virus was performed in Biosafety Level 3 containment using protocols for SARS-CoV-2 approved by the Africa Health Research Institute Biosafety Committee. We used ACE2-expressing H1299-E3 cells for the initial isolation (P1 stock) followed by passaging in Vero E6 cells (P2 and P3 stocks, where P3 stock was used in experiments). ACE2-expressing H1299-E3 cells were seeded at 1.5 × 10^5^ cells per mL and incubated for 18–20 h. After one DPBS wash, the subconfluent cell monolayer was inoculated with 500 μL universal transport medium diluted 1:1 with growth medium filtered through a 0.45-μm filter. Cells were incubated for 1 h. Wells were then filled with 3 mL complete growth medium. After 8 days of infection, cells were trypsinized, centrifuged at 300 rcf for 3 min and resuspended in 4 mL growth medium. Then 1 mL was added to Vero E6 cells that had been seeded at 2 × 10^5^ cells per mL 18–20 h earlier in a T25 flask (approximately 1:8 donor-to-target cell dilution ratio) for cell-to-cell infection. The coculture of ACE2-expressing H1299-E3 and Vero E6 cells was incubated for 1 h and the flask was then filled with 7 mL of complete growth medium and incubated for 6 days. The viral supernatant (P2 stock) was aliquoted and stored at −80 °C and further passaged in Vero E6 cells to obtain the P3 stock used in experiments as follows: a T25 flask (Corning) was seeded with Vero E6 cells at 2 × 10^5^ cells per mL and incubated for 18–20 h. After one DPBS wash, the subconfluent cell monolayer was inoculated with 500 μL universal transport medium diluted 1:1 with growth medium and filtered through a 0.45-μm filter. Cells were incubated for 1 h. The flask was then filled with 7 mL of complete growth medium. After infection for 4 days, supernatants of the infected culture were collected, centrifuged at 300 rcf for 3 min to remove cell debris and filtered using a 0.45-μm filter. Viral supernatant was aliquoted and stored at −80 °C.

### Microneutralization using the focus-forming assay

The Delta variant virus was isolated as previously described (13). Vero E6 cells were plated in a 96-well plate (Corning) at 30,000 cells per well 1 day before infection. Approximately 5 mL sterile water was added between wells to prevent wells at the edge drying more rapidly, which we have observed to cause edge effects resulting in lower number of foci. Plasma was separated from EDTA-anticoagulated blood by centrifugation at 500 rcf for 10 min and stored at −80 °C. Aliquots of plasma samples were heat-inactivated at 56 °C for 30 min and clarified by centrifugation at 10,000 rcf for 5 min, after which the clear middle layer was used for experiments. Inactivated plasma was stored in single-use aliquots to prevent freeze–thaw cycles. For experiments, plasma was serially diluted and the the GenScript A02051 anti-spike monoclonal antibody was added as a positive control to one column of wells. Final plasma dilutions used were 1:25, 1:50, 1:100, 1:200, 1:400, 1:800, 1:1600 for all plasma samples tested. Virus stocks were used at approximately 50-100 focus-forming units per microwell and added to diluted plasma; antibody– virus mixtures were incubated for 1 h at 37 °C, 5% CO_2_. Cells were infected with 100 μL of the virus–antibody mixtures for 1 h, to allow adsorption of virus. Subsequently, 100 μL of a 1X RPMI 1640 (Sigma-Aldrich, R6504), 1.5% carboxymethylcellulose (Sigma-Aldrich, C4888) overlay was added to the wells without removing the inoculum. Cells were fixed at 18 h after infection using 4% paraformaldehyde (Sigma-Aldrich) for 20 min. For staining of foci, a rabbit anti-spike monoclonal antibody (BS-R2B12, GenScript A02058) was used at 0.5 μg/mL as the primary detection antibody. Antibody was resuspended in a permeabilization buffer containing 0.1% saponin (Sigma-Aldrich), 0.1% BSA (Sigma-Aldrich) and 0.05% Tween-20 (Sigma-Aldrich) in PBS. Plates were incubated with primary antibody overnight at 4 °C, then washed with wash buffer containing 0.05% Tween-20 in PBS. Secondary goat anti-rabbit horseradish peroxidase (Abcam ab205718) antibody was added at 1 μg/mL and incubated for 2 h at room temperature with shaking. The TrueBlue peroxidase substrate (SeraCare 5510-0030) was then added at 50 μL per well and incubated for 20 min at room temperature. Plates were then dried for 2 h and imaged using a Metamorph-controlled Nikon TiE motorized microscope with a 2X objective or ELISPOT instrument with built-in image analysis (C.T.L). For microscopy images, automated image analysis was performed using a custom script in MATLAB v.2019b (Mathworks), in which focus detection was automated and did not involve user curation.

### Multi-epitope protein microarray

ImmuSAFE COVID-19 Array slides (Sengenics Corporation, Singapore) were used to measure the anti-SARS CoV-2 IgG and IgA antibodies against N and S proteins. The microarray-based assays were performed as previously described (Smith et al 2021) with the following modifications. After blocking, the 24-plex microarrays were immediately incubated with the samples. Heat-inactivated clarified plasma was diluted 1:50 directly in assay buffer (PBST, 0.1% BSA, 0.1% milk powder) and further processed as described (Smith et al 2021). Wells were washed individually three times with 150µl PBST (PBS, 0.2% Tween-20, pH 7.4). Gaskets were then removed, and arrays washed 2 times with 3ml PBST and twice with 3ml PBS. Arrays were then incubated simultaneously with two detection antibodies: AF647 anti-human IgG and AF555 anti-human IgA (ThermoFisher; 1.25µg/ml each in 3ml assay buffer) for 30 min at RT with gentle agitation and dried by centrifugation at 1200× *g* for 2 min. Arrays were then scanned using the InnoScan 710 (Agilent, Santa Clara, CA, USA) fluorescence microarray scanner to generate 16-bit TIFF files. A GAL (GenePix Array List) file containing the identities of each spot was used to extract data from the TIFF images. Data quantification and extraction was automated using Mapix Software (v8.5.0; Innopsys, after which data was analyzed in R Studio. Briefly, for each spot, the neighbourhood background intensity was subtracted from the foreground intensity. As a threshold, the mean plus 2 standard deviations of the background was applied, and negative values zeroed before the mean of triplicate replicate spots was calculated. Any antigens with a co-efficient of variance (CV) > 20% for the technical replicates were flagged, the outlier spots removed and CVs recalculated based on the two remaining spots per antigen. Reciprocal titres were calculated as previously described (Smith et al, 2021). The cumulative tire of the N protein N-terminal domain, C-terminal domain, epitope 1, epitope 2 and epitope 3 was obtained. The thresholds for S and N proteins were determined using the pre-pandemic controls; for the S proteins the mean plus 2x standard deviation of the pre-pandemics were used to set the threshold, whereas for the N proteins the Optimal Cutpoints R package was used, with an emphasis of maximising the specificity (Lopez-Raton et al, 2013).

### Statistics and fitting

All statistics and fitting were performed using MATLAB v.2019b. Neutralization data were fit to Tx=1/1+(D/ID_50_).

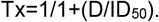

Here Tx is the number of foci normalized to the number of foci in the absence of plasma on the same plate at dilution D and ID_50_ is the plasma dilution giving 50% neutralization. PRNT_50_ = 1/ID_50_. Values of PRNT_50_ <1 are set to 1 (undiluted), the lowest measurable value.

## Data Availability

All data are available in the manuscript or upon reasonable request.

## Acknowledgements

This study was supported by a South African Medical Research Council Award (6084COAP2020) and the Bill and Melinda Gates award INV-018944 to AS. DA was funded through the SAMRC Self-Initiated Grant and the NRF of South Africa Thuthuka (grant#TTK160517165310), the NRF Research Career Advancement Fellowship (grant#RCA13101656388), and a European & Developing Countries Clinical Trials Partnership (EDCTP) senior fellowship (grant#TMA2017SF-1960). JMB thanks the National Research Foundation for a SARChI grant.

## COMMIT-KZN Team

Adrie Steyn, University of Alabama at Birmingham, Africa Health Research Institute.

Alasdair Leslie, Africa Health Research Institute and Division of Infection and Immunity, University College London.

Dirhona Ramjit, Africa Health Research Institute.

Emily Wong, Africa Health Research Institute and Division of Infectious Diseases, University of Alabama at Birmingham.

Guy Harling, Africa Health Research Institute and the Institute for Global Health, University College London, London, UK.

Henrik Kloverpris, Africa Health Research Institute, Division of Infection and Immunity, University College London and Department of Immunology and Microbiology, University of Copenhagen.

Jackson Marakalala, Africa Health Research Institute.

Janet Seeley, Africa Health Research Institute.

Jennifer Giandhari, Kwazulu-Natal Research Innovation and Sequencing Platform.

Kaylesh Dullabh, Department of Cardiothoracic Surgery, University of KwaZulu-Natal, Durban, South Africa.

Kennedy Nyamande, Department of Pulmonology and Critical Care, University of KwaZulu-Natal, Durban, South Africa.

Kobus Herbst, Africa Health Research Institute and the South African Population Research Infrastructure Network, Durban, South Africa

Kogie Naidoo, Centre for Aids Programme of Research in South Africa, University of KwaZulu-Natal

Matilda Mazibuko, Africa Health Research Institute

Moherndran Archary, Department of Paediatrics and Child Health, University of KwaZulu-Natal, Durban, South Africa.

Mosa Moshabela, College of Health Sciences, University of KwaZulu-Natal, Durban, South Africa.

Nesri Padayatchi, Centre for Aids Programme of Research in South Africa, University of KwaZulu-Natal.

Nigel Klein, Africa Health Research Institute and the Institute of Child Health, University College London, London, UK.

Nikiwe Mbatha, Africa Health Research Institute.

Nokuthula Ngcobo, Africa Health Research Institute.

Nokwanda Gumede, Africa Health Research Institute.

Nokwanda Ngcobo, Africa Health Research Institute.

Philip Goulder, Africa Health Research Institute and Department of Paediatrics, Oxford, UK.

Prakash Jeena, Department of Paediatrics and Child Health, University of KwaZulu-Natal, Durban, South Africa.

Rajhmun Madansein, Department of Cardiothoracic Surgery, University of KwaZulu-Natal, Durban, South Africa.

Ravindra K. Gupta, Africa Health Research Institute and Cambridge Institute of Therapeutic Immunology & Infectious Disease, Cambridge, UK.

Rohen Harrichandparsad, Department of Neurosurgery, University of KwaZulu-Natal, Durban, South Africa.

Samita Singh, Africa Health Research Institute.

Thandeka Khoza, Africa Health Research Institute.

Theresa Smit, Africa Health Research Institute.

Thumbi Ndung’u, Africa Health Research Institute, Division of Infection and Immunity University College London, HIV Pathogenesis Programme, The Doris Duke Medical Research Institute and Max Planck Institute for Infection Biology, Berlin.

Vinod Patel, Department of Neurology, University of KwaZulu-Natal, Durban, South Africa.

Zaza Ndhlovu, Africa Health Research Institute.

---

**Fig S 1:**
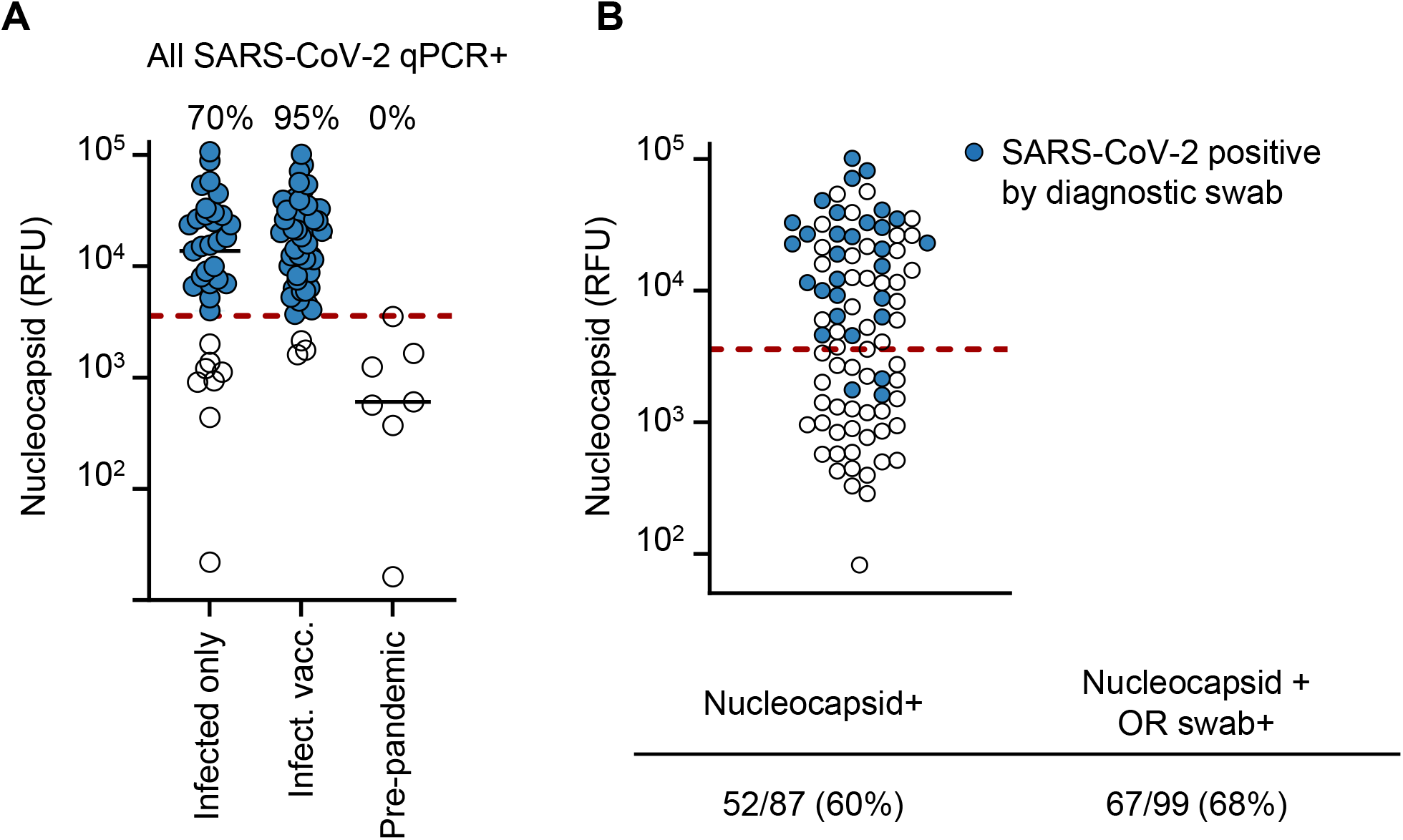
Determination of previous exposure to SARS-CoV-2 by presence of antibodies to SARS-CoV-2 nucleocapsid. (A) Validation of approach. Blood from 90 infected unvaccinated or infected vaccinated participants who were confirmed SARS-CoV-2 infected by qPCR of a nasopharyngeal/oropharyngeal swab and 6 pre-pandemic controls were tested for antibodies to nucleocapsid (Materials and methods). 27 out of 35 (70%) of infected only and 52 out of 55 (95%) of infected and Ad26.CoV2.S vaccinated swab confirmed participants were nucleocapsid antibody positive. All pre-pandemic controls were negative (0/6). (B) Fraction of vaccinated HCW participants with detectable nucleocapsid. Nucleocapsid antibody detection was performed for 87 out of 99 vaccinated participants, with the remaining 12 participants being confirmed for SARS-CoV-2 exposure by qPCR and not included in the nucleocapsid assay. 52 out of 87 (60%) of vaccinated HCW tested were nucleocapsid positive. When the remaining 12 HCW confirmed for SARS-CoV-2 by qPCR were added, the exposure rate increased to 67 out of 99 (68%). Dashed lines in (A) and (B) denote threshold for nucleocapsid detection (Materials and methods). Blue points in (B) denote participants who tested SARS-CoV-2 positive by qPCR.

**Fig S 2:**
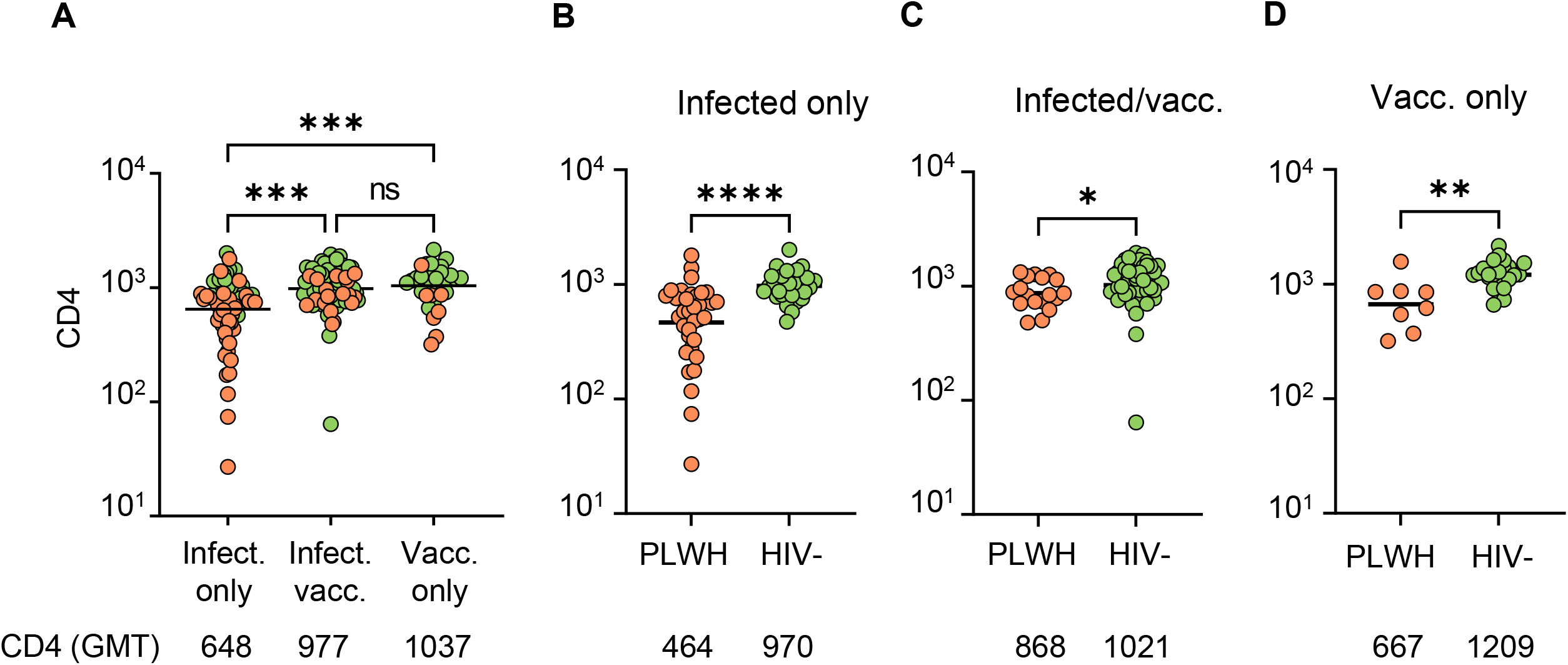
CD4 T cell counts in PLWH and HIV-uninfected participants in the different groups. (A) Comparison of CD4 counts across groups. (B-D) Comparison of CD4 counts between PLWH and HIV-uninfected participants in the infected unvaccinated (B), infected and vaccinated (C), and vaccinated only (D) groups. The CD4 GMT value is list below each group. p-values are * <0.05, ** <0.01, *** < 0.001, **** < 0.0001 as determined by the Kruskal-Wallis test with Dunn multiple hypothesis correction (A) or Mann–Whitney U test (B-D).

**Fig S 3:**
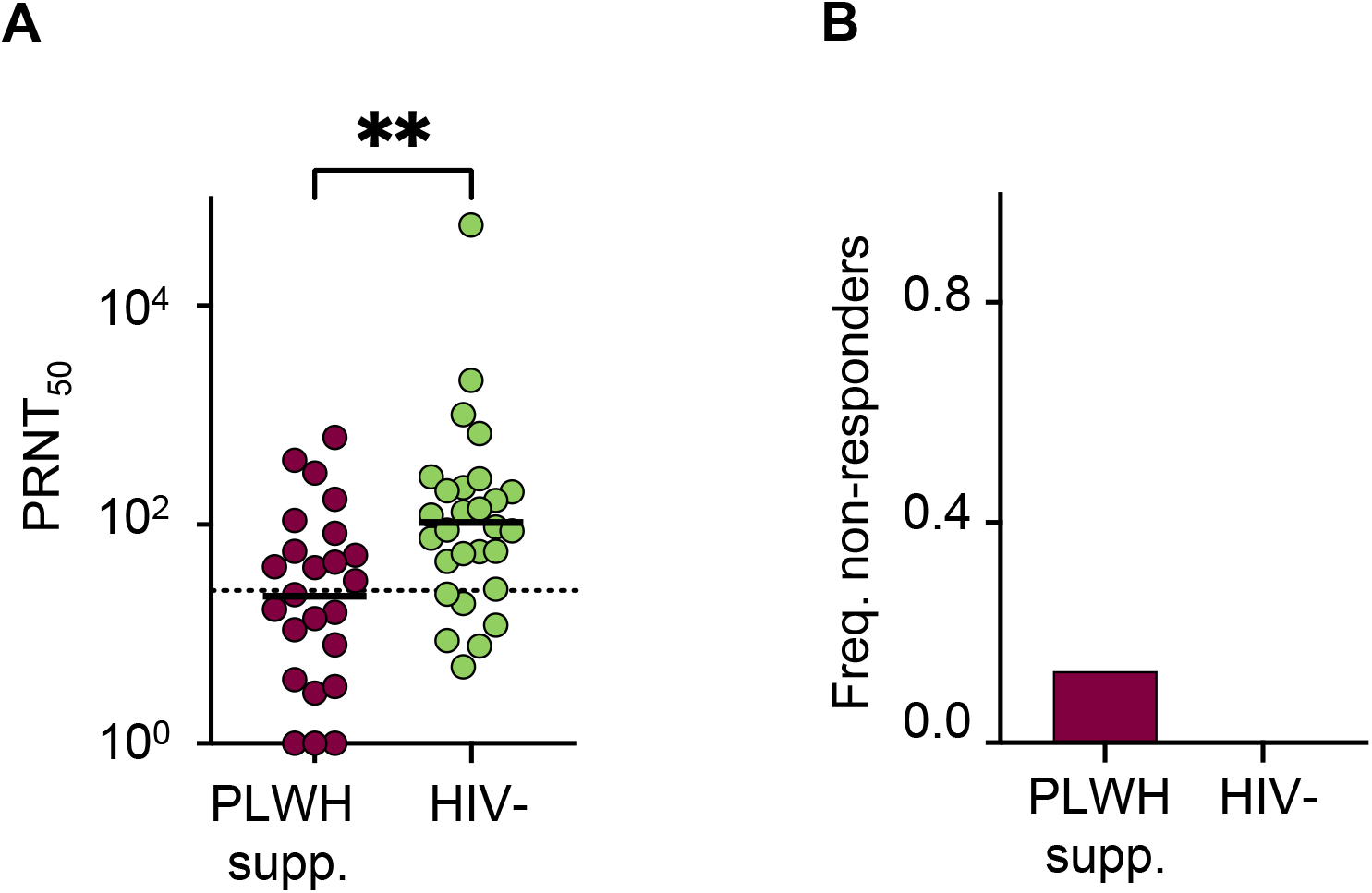
Differences in Delta neutralization between HIV suppressed and HIV-uninfected participants. (A) Neutralization capacity as PRNT50 for Delta variant neutralization in SARS-CoV-2 infected unvaccinated particiupants who are either HIV suppressed (purple points) or HIV uninfected (green points). (B) Frequency of non-responders in (A). p-values are p=0.0028, as determined for (A) by the Mann–Whitney U test and p=0.092 for (B) by Fisher’s exact test.

**TABLE S1:** Characteristics of pre-pandemic participants

|  | All<br>(n=10) | HIV-<br>(n= 5, 50%) | HIV+<br>(n=5, 50%) |
| --- | --- | --- | --- |
| Age years | 37 (24 - 43) | 43 (34 - 44) | 24 (24 - 40) |
| Male sex | 2 (20.0%) | 2 (40.0%) | 0 (0.0%) |
| Fraction HIV viremic | - | - | 3 (60.0%) |
| CD4 cells/μL | 666 (398 - 1014) | 1046 (853 – 1246.5) | 398 (363 - 491) |

All values are median (IQR). Fraction HIV viremic is number PLWH with HIV RNA >40 copies/ml of total PLWH.

